# Accuracy of Healthcare Professionals’ Nasopharyngeal Swab Technique in SARS-CoV-2 Specimen Collection

**DOI:** 10.1101/2020.10.19.20213140

**Authors:** Robbie S. R. Woods, Kellie Nwaokorie, Jana Crowley, Michael Walsh, Eoghan de Barra, Peter D. Lacy

**Author notes:** Author responsible for correspondence and proofs, Robbie Woods, Department of Otolaryngology, Beaumont Hospital, Dublin, Ireland.

## Abstract

**Background:** The COVID-19 pandemic has caused huge pressure on healthcare systems worldwide. Public health measures to control the virus are reliant on testing, including appropriate collection of specimens for analysis.

**Methods:** A prospective study of nasopharyngeal swab technique by staff in an academic tertiary referral centre was carried out. Nasopharyngeal swab technique was evaluated by a novel design of a navigated swab on a three-dimensional model head.

**Results:** Swab technique of 228 participants was assessed. Technique was poor, with a success rate of nasopharyngeal swabbing at 38.6%. Angle and length of insertion were significantly different between those with successful and unsuccessful technique. Doctors were significantly more accurate than nurses and non-healthcare professionals (p<0.01).

**Conclusion:** Inaccurate specimen collection from poor swab technique could contribute to a false negative rate of testing for SARS-CoV-2. Specific training in nasopharyngeal anatomy and swab technique may improve the accuracy of nasopharyngeal swabbing.

## Introduction

The outbreak of a novel coronavirus, designated SARS-CoV-2, and the consequent respiratory illness COVID-19 has caused a profound impact on healthcare systems worldwide.^1^ Reliable laboratory diagnosis is a critical component of public health interventions to control the COVID-19 pandemic.^2^ Testing for respiratory tract viral RNA with real-time polymerase chain reaction (PCR) has been the mainstay in diagnosis.^3^ This testing is contingent on viral RNA being present in the sample collected.^4^

It is likely that viral detection rates from samples of different sites, such as the nasopharynx, oropharynx or lower respiratory tract, will vary over the course of the illness and from patient to patient.^4^ However, current evidence suggests a higher viral load is present in the nasopharynx than the oropharynx in COVID-19.^5^

The sensitivity of PCR from nasopharyngeal specimens is poor, ranging from 52-71%.^6^ Although PCR generally has a very high specificity, sensitivity is less certain and can depend on the targets used and variability of the viral genome.^7^ Frequent sequencing to identify mutations and adjust primers and probes may alleviate this risk.^4^ Other possible explanations for false-negative specimens include a viral load below the detectable limit of the assay, diminished upper airway viral shedding as the disease progresses and suboptimal collection or handling technique.^6^

Correct sampling technique is important to ensure the accuracy of the test is not affected by the quality of the sample. Nasopharyngeal sample quality can be greatly affected by precise location, pressure, direction, and number of swab strokes employed. Advice on sampling for SARS-CoV-2 from the nasopharynx has been published.^8, 9^ A study was performed to assess nasopharyngeal swab technique of staff in a major academic institution.

## Materials and methods

A prospective study of swab technique among hospital staff was carried out over two consecutive days. All healthcare staff at the institution were invited to participate and informed consent for participation was obtained. The study was approved by the hospital ethics committee. The authors assert that all procedures contributing to this work comply with the ethical standards of the Beaumont Hospital ethics committee and with the Helsinki Declaration of 1975, as revised in 2008. Participants were asked whether they had taken a swab for SARS-CoV-2 in the clinical setting and if they had received formal training in technique. Participants then performed a nasal swab using a three-dimensional navigated swab on a life-size model head.

A validated PHACON Sinus Trainer (Leipzig, Germany) with a cassette containing normal sinus anatomy was used as a model.^10^ This was placed at a 70-degree angle, confirmed by goniometer, to represent a typical patient sitting for a swab in their car. The nose was positioned at a height of 120 cm. Figure 1 demonstrates the arrangement of the model. A navigated swab was created by inserting a tip-tracked electromagnetic navigable stylet (Medtronic, Minneapolis, MN, USA) into a standard nasopharyngeal culture swab and securing it in position with glue, as shown in Figure 2. Using the Medtronic StealthStation® navigation system (Minneapolis, MN, USA) with computed tomography imaging corresponding to the Sinus Trainer, the nasopharynx was marked out in three dimensions with a tumour marking programme. The nasopharynx was measured at 91 mm from the nasal tip. From this point, a space of 21 mm depth, 19 mm height and 30 mm width was demarcated, to correlate with measurements in the literature.^11^ This was used to represent the position of accurate nasopharyngeal sampling, as shown in the red area marked in Figure 3.

**Figure 1:**
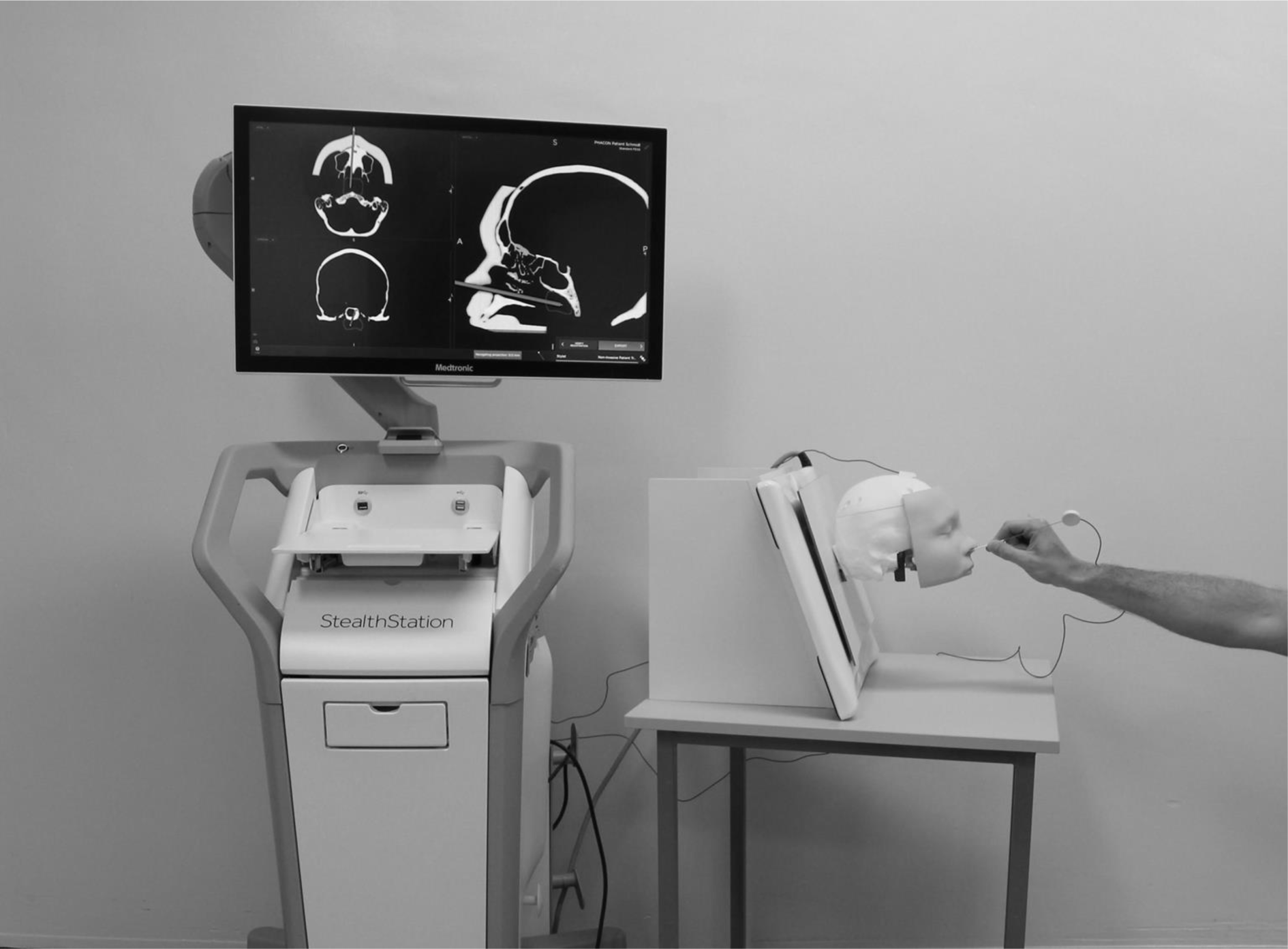
Model for Assessment of Nasopharyngeal Swab Technique

**Figure 2:**
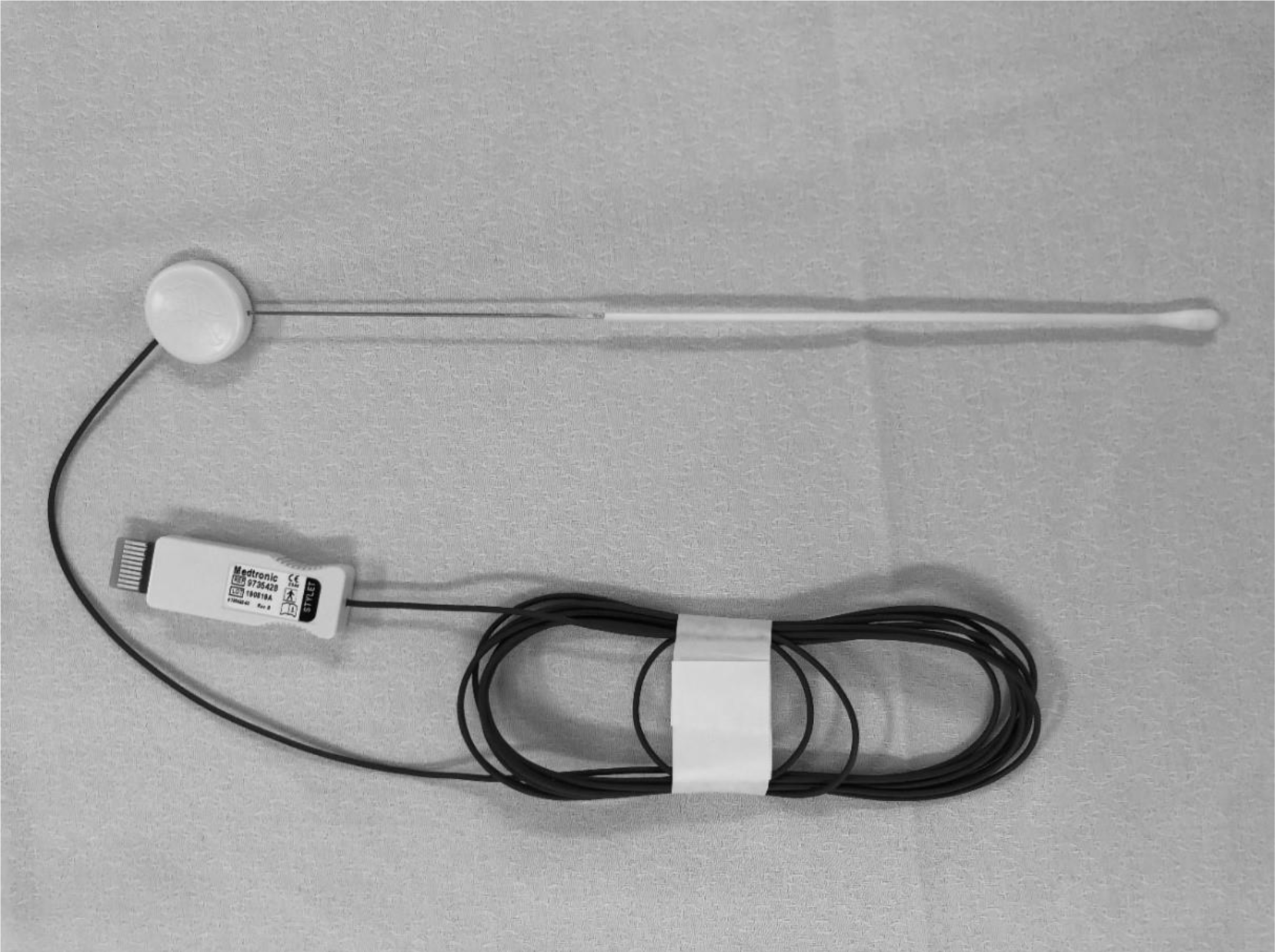
Navigated Swab Design

**Figure 3:**
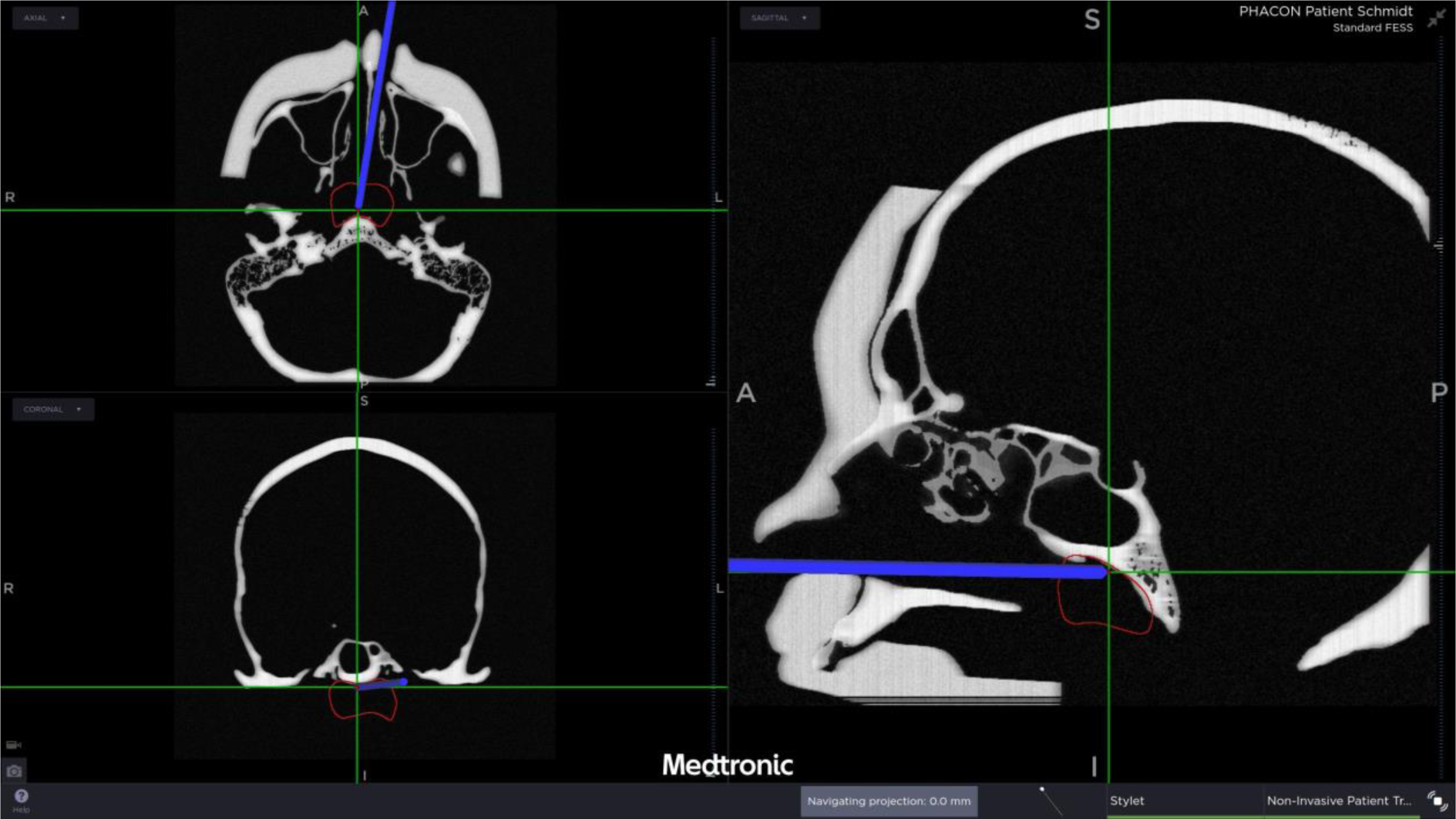
Three-Dimensional Navigation of Swab with Nasopharynx Marked in Red

All participants were asked to swab as they would expect to for a patient with suspected SARS-CoV-2, pausing in position where the sample would be collected, whereupon a navigation screenshot was captured. The navigation screen was not visible to the participant. Results were recorded as a successful swab if present in the marked nasopharynx, as shown in Figure 3. Metadata from the navigation system was analysed to ensure no discrepancies.

ImageJ software (National Institutes of Health, Maryland, United States) was used to analyse swab position.

Statistical analysis was performed using IBM SPSS Statistics version 23.0.0.2 software (IBMCorp. NewYork, United States). Significance of association tests between swab technique and subgroups were performed using the two-tailed Fisher Exact test for independence. Testing for association with continuous variables was performed using the T test, if normally distributed, or the Mann-Whitney U test, if not normally distributed. The threshold for statistical significance was set at p≤0.05, and tests were two-sided.

## Results and analysis

The study had 230 participants. Two participants were excluded due to the incorrect gathering of data, leaving a total of 228 participants. Characteristics of participants are summarized in Table I. Seniority within roles is shown in Table II. Other healthcare professionals included dieticians, occupational therapists, physician associates, physiotherapists, radiographers and speech and language therapists. Non-healthcare staff included all hospital staff not involved in regular patient contact.

**TABLE I.** Characteristics of Study Participants

| Role | Number of Participants | Male | Female | Previously Performed a Swab | Received Training in Swab Technique |
| --- | --- | --- | --- | --- | --- |
| Nurse | 92 | 12 (13%) | 80 (87%) | 45 (49%) | 35 (38%) |
| Doctor | 81 | 46 (57%) | 35 (43%) | 35 (43%) | 18 (22%) |
| Other Healthcare | 32 | 4 (13%) | 28 (87%) | 5 (16%) | 5 (16%) |
| Non-Healthcare | 23 | 6 (26%) | 17 (74%) | 0 (0%) | 0 (0%) |
| Total | 228 | 68 (30%) | 160 (70%) | 85 (37%) | 58 (25%) |

**TABLE II.** Breakdown of Seniority

| Seniority | Number of Participants |
| --- | --- |
| Nurse Manager | 30 |
| Staff Nurse | 62 |
| Consultant | 7 |
| Registrar | 15 |
| Senior House Officer | 27 |
| Intern | 32 |

The overall success rate in correctly performing a nasopharyngeal swab was 38.6% (88/228). The number of successful swabs by role is shown in Figure 4. Successful swab rates were low amongst all groups including nurses (28/92 (30%)), doctors (45/81 (56%)), other healthcare professionals (12/32 (38%)) and non-healthcare staff (3/23 (13%)). Of the 85 staff members who had previously performed a swab for SARS-CoV-2, only 45 (53%) had received training.

**Figure 4:**
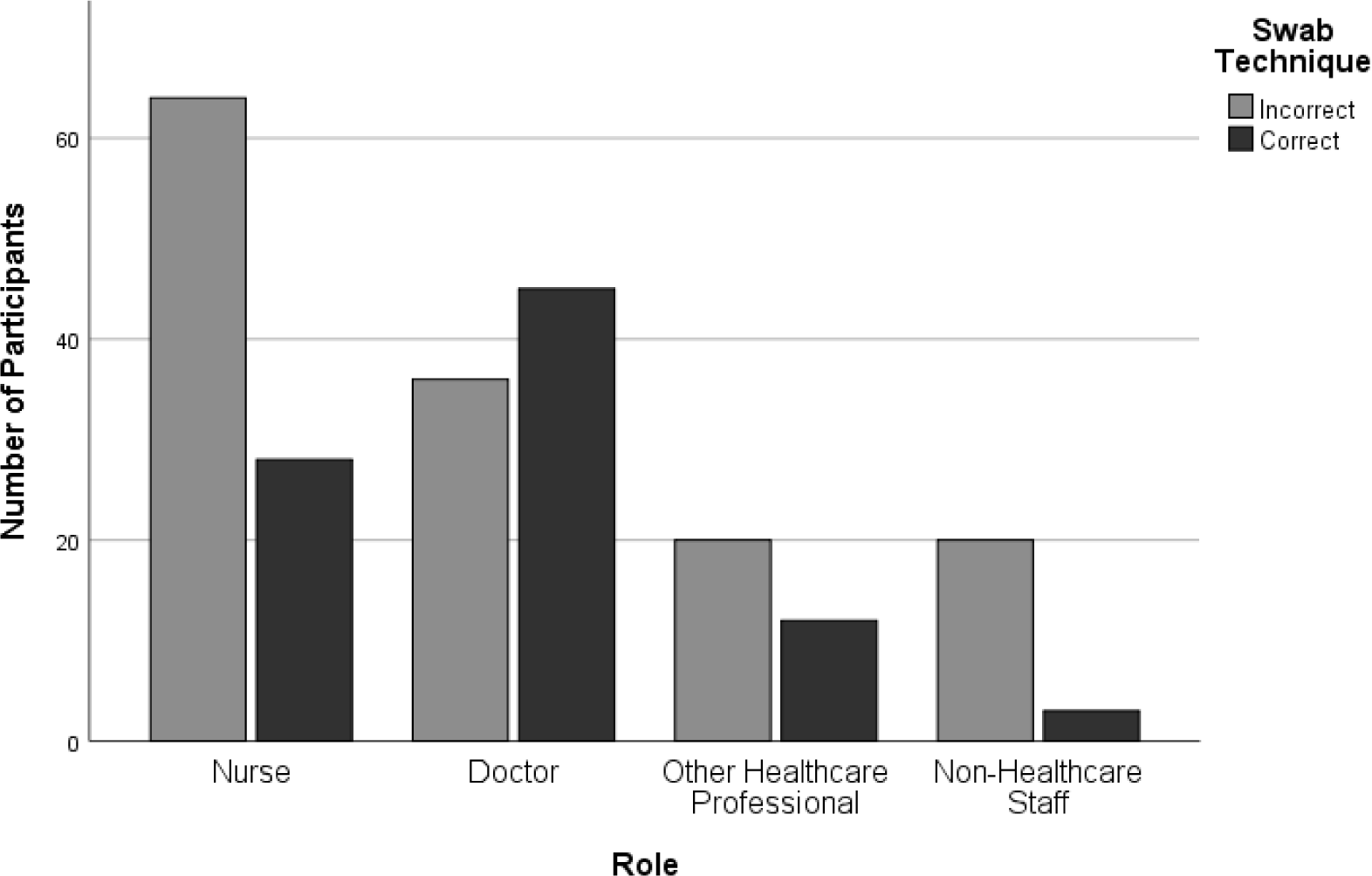
Successful Swab Technique by Role

Median length of insertion was significantly greater in those with successful swab technique at 11.75cm (Interquartile range 1.81cm) than those with unsuccessful technique at 7.86cm (Interquartile range 4.46cm) (p<0.05). Median angle of insertion was significantly shallower in those with successful technique at 0.58 degrees (Interquartile range 0.4 degrees) than those with unsuccessful technique at 25.59 degrees (Interquartile range 16.84 degrees) (p<0.05).

Swab technique was significantly more correct in doctors than nurses (p<0.01) and non-healthcare staff (p<0.01). There was no significant difference between other groups, as shown in Table III. Comparing by other characteristics, as shown in Table 3, swab technique was significantly more accurate when performed by males (p<0.01). Participants who either performed a previous swab or received training in swab technique were not significantly more accurate than those who did not. On performing logistic multivariate forward stepwise regression to eliminate confounders, the only significant factor that determined accurate swab technique was the role of a doctor (odds ratio 0.296, 95% confidence interval 0.137-0.639, p<0.01).

**TABLE III.**
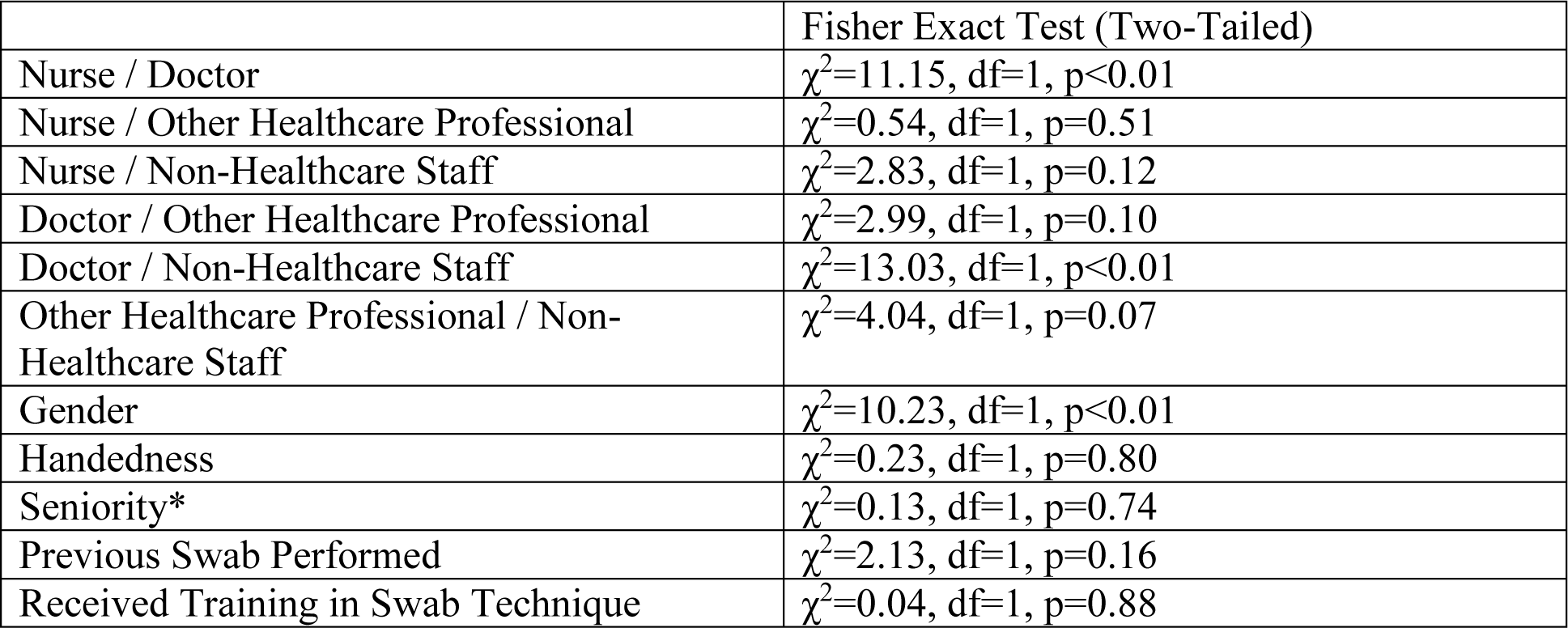
Comparison of Swab Technique Between Roles and Subgroups *Seniority compared Nurse Manager, Consultant and Registrar against Staff Nurse, Senior House Officer and Intern.

|  | Fisher Exact Test (Two-Tailed) |
| --- | --- |
| Nurse / Doctor | $\chi^2=11.15$, df=1, p<0.01 |
| Nurse / Other Healthcare Professional | $\chi^2=0.54$, df=1, p=0.51 |
| Nurse / Non-Healthcare Staff | $\chi^2=2.83$, df=1, p=0.12 |
| Doctor / Other Healthcare Professional | $\chi^2=2.99$, df=1, p=0.10 |
| Doctor / Non-Healthcare Staff | $\chi^2=13.03$, df=1, p<0.01 |
| Other Healthcare Professional / Non-Healthcare Staff | $\chi^2=4.04$, df=1, p=0.07 |
| Gender | $\chi^2=10.23$, df=1, p<0.01 |
| Handedness | $\chi^2=0.23$, df=1, p=0.80 |
| Seniority* | $\chi^2=0.13$, df=1, p=0.74 |
| Previous Swab Performed | $\chi^2=2.13$, df=1, p=0.16 |
| Received Training in Swab Technique | $\chi^2=0.04$, df=1, p=0.88 |

## Discussion

Accurate specimen collection is critical to ensure optimal sensitivity of testing for SARS-CoV-2 but depends on the skill of the person performing the swab.^12^ In many cases, healthcare professionals have been redeployed to other departments to provide enough staff for testing, including non-otolaryngologists taking nasopharyngeal swabs.^13^ Appropriate training is particularly important for staff unfamiliar with the relevant anatomy, although training is not always adequate.^13^

As nasopharyngeal swab technique is likely to vary, this study aimed to investigate the techniques being used in this institution. A novel, but easily reproducible, training tool was created using a navigated swab on a three-dimensional model to give immediate feedback on technique to staff. Overall successful swab technique was low at 38.6%, although only 53% of those who had previously performed a swab received training. Prior training in swab technique was not shown to influence the ability to perform a successful swab. The only factor that significantly influenced successful swab technique was the role of doctor. This may be due to a better knowledge of appropriate anatomy,^13^ dedicated training in Otolaryngology at undergraduate level or better understanding of the given task. However, it is also possible that the training received by staff performing swabs at this institution may not be adequate. For example, staff familiar with routine nasal swabbing for other pathogens may not be familiar with the required angle and length of insertion required to reach the nasopharynx.

In the absence of therapeutic agents, crucial to many public health approaches to control the pandemic has been widespread testing.^3^ As there is currently no gold standard for the diagnosis of COVID-19, sensitivity and specificity of PCR from nasopharyngeal swabs are difficult to accurately calculate.^14^ Comparators used have included those with evidence of COVID-19 on imaging and those with a previously positive swab on PCR.^15^ Reported accuracy of testing varies both by institution, with sensitivity in some reported up to 96%,^12^ and by stage of disease, when sensitivity can drop to below 70% after day eight of illness.^16^ Sensitivity of PCR can be improved by increasing targets, rapid testing to prevent degradation of samples and testing larger volume of sample in cases of lower viral load.^17^ However, it is critical to ensure an adequate yield of specimen through appropriate swab technique to maximise the accuracy of PCR. Although alternatives such as exhaled breath condensate or rapid salivary testing are being considered,^18, 19^ nasopharyngeal swab testing remains standard practice in the diagnosis of SARS-CoV-2.

There are some limitations in this study. The technique evaluated considered swab position but did not evaluate duration of swab and number of swab twists performed. Participants were not asked if they had a swab performed on themselves, which may have improved their knowledge of accurate technique. Although an angle of 70 degrees was chosen to reflect clinical practice, alteration of head position in clinical settings may allow improved access to the nasopharynx. Only technique in nasopharyngeal swab was assessed but testing of specimens from multiple sites may improve overall sensitivity.^20^

## Conclusion

This study demonstrates, using a novel training tool, objective evidence of poor technique in nasopharyngeal swabbing, with a rate of successful technique at 38.6%. Universal standardisation of swab technique with adequate training may improve specimen collection and therefore potentially improve sensitivity of testing to maximise the benefits of public health testing programmes.

## Data Availability

Anonymous data available from research team. All data presented.

## Acknowledgements

Garreth O’Callaghan, Ann Aherne and Rory Flood for providing technical support to the project.

## Bullet Point Summary

- There is little evidence on the accuracy of swabbing technique in peer-reviewed published medical literature
- This study uses a novel tool to evaluate a crucial aspect of public health measures to control the spread of SARS-CoV-2
- The low success rate of accurately swabbing the nasopharynx implies that better training is necessary
- Better training may improve specimen collection and sensitivity of testing for SARS-CoV-2
- Standardised training videos with description of the relevant anatomy would likely be useful to improve testing

## References

1. Tanne JH, Hayasaki E, Zastrow M, Pulla P, Smith P, Rada AG. Covid-19: how doctors and healthcare systems are tackling coronavirus worldwide. BMJ 2020;368:m1090

2. Lu X, Wang L, Sakthivel SK, Whitaker B, Murray J, Kamili S, et al. US CDC Real-Time Reverse Transcription PCR Panel for Detection of Severe Acute Respiratory Syndrome Coronavirus 2. Emerg Infect Dis 2020;26

3. Binnicker MJ. Emergence of a Novel Coronavirus Disease (COVID-19) and the Importance of Diagnostic Testing: Why Partnership between Clinical Laboratories, Public Health Agencies, and Industry Is Essential to Control the Outbreak. Clin Chem 2020;66:664–6

4. Patel R, Babady E, Theel ES, Storch GA, Pinsky BA, St George K, et al. Report from the American Society for Microbiology COVID-19 International Summit, 23 March 2020: Value of Diagnostic Testing for SARS-CoV-2/COVID-19. mBio 2020;11

5. Zou L, Ruan F, Huang M, Liang L, Huang H, Hong Z, et al. SARS-CoV-2 Viral Load in Upper Respiratory Specimens of Infected Patients. N Engl J Med 2020;382:1177–9

6. Bullis SSM, Crothers JW, Wayne S, Hale AJ. A Cautionary Tale of False-Negative Nasopharyngeal COVID-19 Testing. IDCases 2020:e00791

7. Lemmon GH, Gardner SN. Predicting the sensitivity and specificity of published real-time PCR assays. Ann Clin Microbiol Antimicrob 2008;7:18

8. Di Maio P, Iocca O, Cavallero A, Giudice M. Performing the nasopharyngeal and oropharyngeal swab for 2019-novel coronavirus (SARS-CoV-2) safely: How to dress, undress, and technical notes. Head Neck 2020

9. Marty FM, Chen K, Verrill KA. How to Obtain a Nasopharyngeal Swab Specimen. N Engl J Med 2020;382:e76

10. Alwani MM, Svenstrup TJ, Bandali EH, Sharma D, Higgins TS, Wu AW, et al. Validity testing of a three-dimensionally printed endoscopic sinonasal surgery simulator. Laryngoscope 2019

11. El-Anwar MW, Ali AH, Elnashar I, Elfiki IM, Ahmed AF, Abdulmonaem G. Normal Nasopharyngeal Measurement by Computed Tomography in Adult. J Craniofac Surg 2017;28:e395–e8

12. Patel ZM. Reflections and new developments within the COVID-19 pandemic. Int Forum Allergy Rhinol 2020;10:587–8

13. Karligkiotis A, Arosio A, Castelnuovo P. How to Obtain a Nasopharyngeal Swab Specimen. N Engl J Med 2020;382

14. Pujadas E, Ibeh N, Hernandez MM, Waluszko A, Sidorenko T, Flores V, et al. Comparison of SARS-CoV-2 detection from nasopharyngeal swab samples by the Roche cobas 6800 SARS-CoV-2 test and a laboratory-developed real-time RT-PCR test. J Med Virol 2020

15. Kokkinakis I, Selby K, Favrat B, Genton B, Cornuz J. [Covid-19 diagnosis : clinical recommendations and performance of nasopharyngeal swab-PCR]. Rev Med Suisse 2020;16:699–701

16. PCR testing for COVID-19: where to swab? : National Centre for Infectious Diseases Agency for Care Effectiveness. Ministry of Health Singapore; 2020; Available from: https://www.moh.gov.sg/docs/librariesprovider5/default-document-library/pcr-testing-for-covid-19---where-to-swab-(24-april-2020).pdf.

17. Torres I, Albert E, Navarro D. Pooling of nasopharyngeal swab specimens for SARS-CoV-2 detection by RT-PCR. J Med Virol 2020

18. Khoubnasabjafari M, Jouyban-Gharamaleki V, Ghanbari R, Jouyban A. Exhaled breath condensate as a potential specimen for diagnosing COVID-19. Bioanalysis 2020

19. Azzi L, Baj A, Alberio T, Lualdi M, Veronesi G, Carcano G, et al. Rapid Salivary Test Suitable for a Mass Screening Program to Detect Sars-Cov-2: A Diagnostic Accuracy Study. J Infect 2020

20. Wang W, Xu Y, Gao R, Lu R, Han K, Wu G, et al. Detection of SARS-CoV-2 in Different Types of Clinical Specimens. JAMA 2020

